# Tracing Alzheimer’s Genetic Footprints: A Pioneering Longitudinal Study Using Artificial Intelligence to Unravel Mutation-Driven Risks and Progression in Virtual Patients; Part 1 – The APOE genotypes

**DOI:** 10.1101/2024.04.02.24305206

**Authors:** WR Danter

## Abstract

Alzheimer’s Disease (AD), the most common neurodegenerative disorder, presents a significant challenge for early detection and intervention due to its complex etiology involving genetic, environmental, and lifestyle factors. This study harnesses the innovative potential of artificial intelligence (AI) through the aiHumanoid platform to simulate AD progression. In this study, we focus on the impact of two APOE genotypes on disease development and progression. Our longitudinal virtual subject simulations, grounded in extensive medical literature and genetic information, explore the nuanced interplay between specific genetic variants, APOE ε3/4 and ε4/4, and their role in AD’s heterogeneity. Despite the potential limitations associated with emerging technologies, including the translatability of AI simulations to real-world scenarios and the scope of genetic variants, this research provides key insights into early biomarkers and the progression patterns of AD. Future segments of this study (Part 2 and Part 3) will broaden the analysis to encompass a wider array of genetic factors and their interactions, enhancing the understanding of AD and paving the way for personalized intervention strategies. Ethical considerations surrounding the use of AI in medical research are acknowledged, emphasizing the need for responsible integration of technology in healthcare. Our findings underscore the transformative potential of AI in advancing AD research, offering a foundation for future studies aimed at refining diagnostic and therapeutic approaches through enhanced realism in simulations and a comprehensive exploration of genetic and environmental factors.

## Introduction

Alzheimer’s Disease (AD), the most prevalent neurodegenerative disorder worldwide, is characterized by a spectrum of symptoms ranging from mild cognitive impairments to severe dementia, leading to death. This diversity in clinical presentations not only diminishes cognitive functions but also significantly impacts the quality of life, emphasizing the need for nuanced care and management across all disease stages [1]. Late-Onset Alzheimer’s Disease (LOAD), which typically emerges after the age of 65, is particularly notable for its complex etiology involving genetic, environmental, and lifestyle factors, necessitating a comprehensive approach to research and treatment.

Among the genetic factors, the Apolipoprotein E (APOE) gene, and specifically its ε4 allele, has been identified as a significant risk factor for LOAD [2, 3]. Individuals carrying one ε4 allele (APOE ε3/4) face an increased risk, while those with two ε4 alleles (APOE ε4/4) are at an even higher risk. The role of APOE in lipid metabolism and its association with amyloid-beta deposition, a hallmark of AD pathology, underscores the importance of targeted research and intervention strategies based on genetic risk profiles [4, 5, 6].

Recent advancements highlight the pivotal role of genetic predispositions in understanding AD, with the APOE ε4 allele being the focal point of LOAD research. Despite extensive studies, the precise mechanisms through which APOE ε4 contributes to LOAD pathogenesis, including aspects like amyloid-beta pathology, tau phosphorylation, and synaptic function, remain only partially understood while the protective role of two copies of APOE ε2 allele has been established. However, these studies pave the way for potential therapeutic strategies targeting APOE ε4, emphasizing its critical role in the search for effective AD treatments [4, 7, 8].

This paper aims to provide an integrated overview of the current understanding of the influence of APOE genotypes on LOAD progression, highlighting recent advancements in the field. By examining the impact of APOE ε3/4 and ε4/4 genotypes and exploring potential interventions, this research contributes to a more nuanced understanding of the disease, facilitating the development of diagnostic tools and therapeutic interventions tailored to individual risk profiles[9.10]. The protective role for the APOE ε2/2 genotype is not addressed in the current study. Similarly, a recently identified rare protective APOE ε4 variant (R251G) serves to emphasize the diverse effects of this gene but is also not considered in this project [11].

Advancing the foundations of our current knowledge [12], this AI-guided longitudinal virtual study seeks to identify potential early biomarkers and delineate distinct progression patterns of AD. By simulating a broad spectrum of genetic mutations, our research aims to unveil phenotypic and genotypic features that may indicate susceptibility to AD at much earlier stages than previously identified, offering a potential revolution in early diagnostic and risk assessment strategies.

The incorporation of AI and virtual patient profiles through the aiHumanoid platform represents a novel methodological leap in the study of Alzheimer’s Disease, promising to enhance our understanding of its genetic underpinnings and progression. Preliminary simulations have highlighted novel biomarkers and neural network changes in virtual patients with specific genotypes, suggesting pathways for early intervention based on blood-based biomarkers [13]. The present paper presents the methodology, implementation, and initial findings of our longitudinal study, emphasizing the transformative potential of AI in the early detection and management of Alzheimer’s Disease. By advancing our knowledge of genetic risk factors and their impact across the lifespan, we strive to pave the way for personalized treatment approaches, ultimately aiming to improve outcomes for individuals at risk of this debilitating disease [12, 14].

The current research project advances our previous simulation of the APOE ε4 variant in the first AI based simulation of a whole brain organoid (aiWBO) [15]. An advanced version of the aiWBO developed in that project forms the CNS organoid component of the current aiHumanoid simulations (v8.3).

## Methods

### Overview

Building upon our pioneering work with artificially induced human stem cells (aiPSC) and organoids (aiOrganoid), this study utilizes aiHumanoid simulations [16,17,18] as subjects in a virtual longitudinal investigation into Alzheimer’s Disease (AD). We aim to better understand the influence of five genetic mutations commonly associated with AD, focusing on early diagnosis and disease progression.

1. Updating the aiHumanoid Simulation to v8.3: The previous version 8.2 of the aiHumanoid [18] underwent modest updates to v8.3. The main differences are that the revised version integrates new simulations for specific AD associated mutations and important dementia subsystems including a new diagnostic subsystem for the diagnosis of AD (Dx of AD). The number of integrated organoid simulations remains at 21. A literature validation of the AD simulations, employing the same approach used in previous versions was applied to the updated simulations comprising v8.3.
2. AD Validation Profile in the aiHumanoid Simulations: To confirm the presence of AD in the affected aiHumanoid simulations, a list of 28 phenotypic features was assembled from the literature for evaluation and are presented in Appendix A. All features were statistically significantly different from controls for multiple age matched cohorts and regarding the diagnosis of AD. This analysis is based on the Wilcoxon signed rank test and the Cliff’s delta effect size estimates.
3. Study Design and Objectives: This is the first of its kind virtual longitudinal study using the aiHumanoid simulations. The objectives of the study were (i) to evaluate the impact of two of the most common, genetic variants associated with the risk of developing Alzheimer’s disease (AD) in the context of aging. (ii) to evaluate a panel of features for the purpose of identifying opportunities for early diagnosis, and (iii) to better understand disease progression from childhood to middle age and beyond.

### The Virtual Subjects used in this study

The profiles for 25 unique and healthy virtual subjects were synthesized by GPT-4, OpenAI’s advanced language model (September 2023 version) https://chat.openai.com/. GPT4 used its extensive training data encompassing medical literature, patient profiles, and related clinical information, to synthesize diverse and representative profiles for 25 healthy subjects. Each subject profile was reviewed by an experienced physician prior to enrolment. This longitudinal study design permitted us to create a virtual study with 3 genotype groups (WT, APOE ε3/4 and APOE ε4/4) X 16 (age groups) X 25 (virtual subjects), the equivalent of data from 1200 patients. The virtual subjects used in this study serve as hypothetical, but commonly encountered examples of risks associated with AD in specific affected cohorts but do not represent actual individuals or precise medical histories.

(Note: GPT-4, developed by OpenAI, is a state-of-the-art language model capable of generating humanlike text based on extensive training data. In this study, it was employed to generate virtual subject profiles, harnessing its vast knowledge from medical literature to ensure the profiles’ accuracy and relevance.)

Inclusion Criteria (Healthy cohorts):

1. Generally healthy at age 5 years
2. Ages 5 years to 80 years
3. Approximately equal representation of males and females
4. All required individual data is available

Exclusion criteria:

5. Age greater than 80 years of age
6. Any documented preexisting genetic abnormalities
7. Any documented disease processes prior to age 5 years

The Affected Subjects:

The LOAD associated genetic variations studied included: the APOE ε3/4 and APOE ε4/4, genotypes. The age cohorts of 25 healthy subjects each underwent AI gene editing to introduce gain of function (GOF) mutations for each of the two AD associated genotypes studied (19). This process created 32 highly matched cohorts where the only difference was the presence or absence of a specific genotype. In these well-matched cohorts, properties like obesity, hypertension and Type 2 DM are essentially emergent properties primarily associated with age. The virtual approach has the major advantage that all patient data was available for analysis since there was no attrition which would be common in traditional longitudinal study of this kind. The data from all cohorts were evaluated beginning at age 5 years and continuing at 5-year intervals up to and including age 80 years of age (16 age cohorts).

Statistical Analysis:

The Null hypothesis states that there are no statistically significant differences or at least medium effect sizes for the two mutated genotypes compared to the age matched healthy subjects. The data was largely not normally distributed, so the non-parametric version of the paired t-test was used, namely the Wilcoxon signed rank test. Given that there many tests (N=28) were conducted, the conservative Bonferroni correction was applied. The corrected p value to achieve significance therefore became 0.05/28 = 0.0019 for this study.

The alternative hypothesis states that there are significant differences in the affected groups compared to the healthy controls.

The true effect size was estimated using Cliff’s delta. Cliff’s delta was used because the data were not normally distributed with a sample size of 25 subjects per cohort. To calculate Cliff’s delta the continuous data was transformed into interval data based on whether the data from the affected group was larger or smaller than the unaffected group. The cliff’s delta was then calculated as (N (larger than) – N (smaller than))/the standard deviation of the differences between the groups (20,21). This produced a range of effect size estimates between -1 (a negative effect) and +1 (a positive effect). A value close to 0 was interpreted as having no effect. To determine the size of the effect we used the following heuristic scale: d <0.147 (negligible), d = 0.147 to <0.330 (small), d = 0.330 to <0.474 (medium) and d >= 0.474(large) as suggested in (22). To compensate for the modest sample size per cohort, all initial Cliff’s d values were modified using the Hedges correction (23) which was calculated to be 0.984. Final effect size estimates were obtained by multiplying the initial effect sizes by 0.984. The 95% CI around the effect size estimate was calculated using the SE of the differences/square root of the sample size (N pairs).

## Results

In the present study, v8.3 (2024) of the DeepNEU database was used, which is characterized by modest upgrades to its previous version v8.2, [18]. Specifically, v8.2 had 7347 concepts and 68148 nonzero causal relationships. Version 8.3 has 7447 concepts and 69581 relationships. This means that for every nonzero concept in the causal relationship matrix, there were more than 9.3 incoming and outgoing causal relationships.

Furthermore, v8.3 offers new validated simulations for the diagnosis of Alzheimer’s disease (AD) and dementia related pathways. The total number of integrated organoid simulations remains unchanged at 21. The previously implemented early stopping algorithm (v8.2) was retained in this updated version. For the purposes of this longitudinal study, we chose to use the results after 14 iterations, as determined through a 3 valued moving average, which minimized any evidence of overfitting.

### Results for APOE ε3/4 Genotype vs Controls (see graphic summary in Table 1.)

APOE ε3/4 Genotype: Alterations in Cognitive and Functional Metrics Across Age Span Reflect Potential AD Progression Markers

**Table 1:**
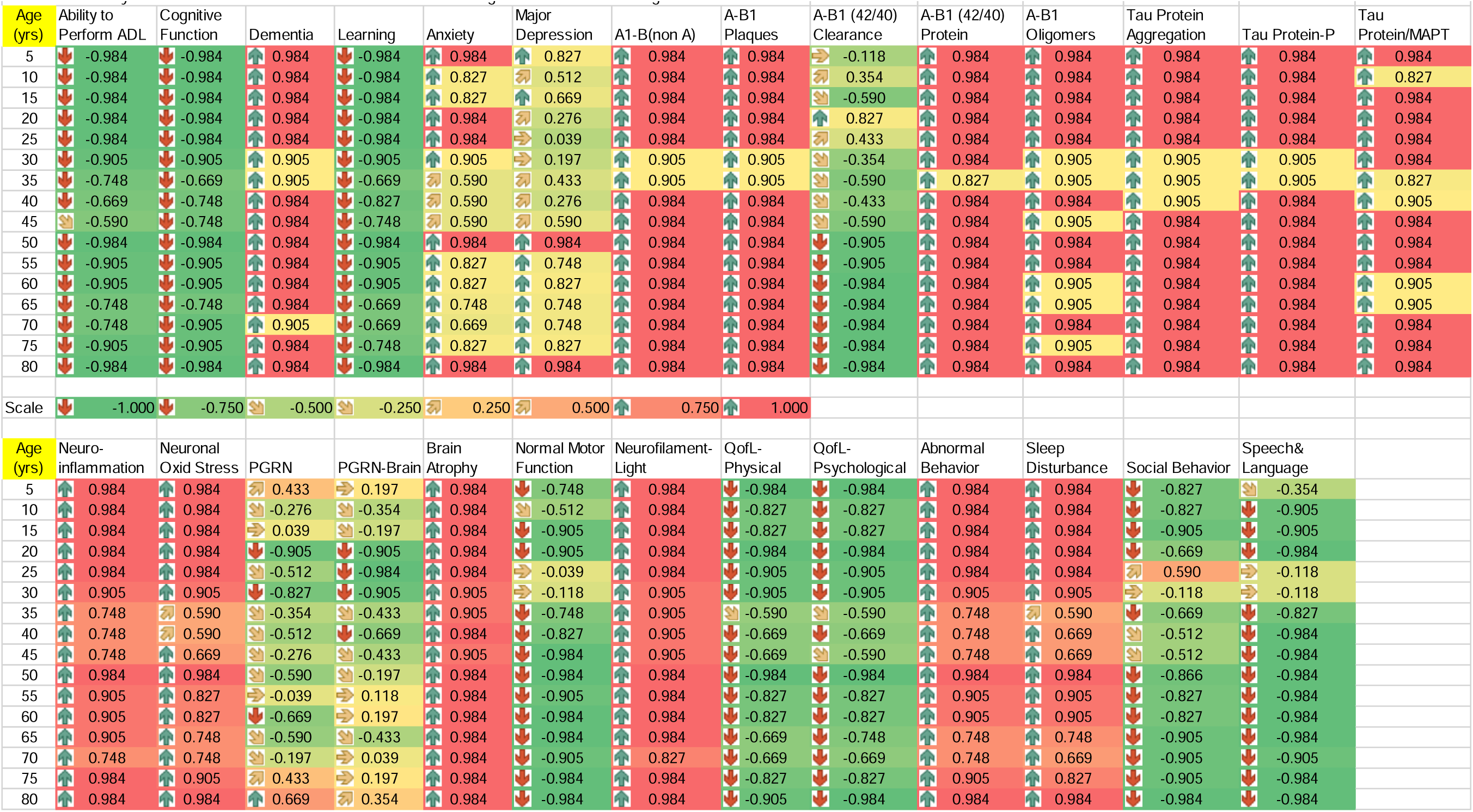
Summary of the Cliff’s delta effect size data for the APOE ε3/4 gene variant across all Age cohorts.

### Statistical Examination of Longitudinal Changes in AD-Related Features

The longitudinal trajectory of Alzheimer’s Disease (AD)-related cognitive and functional attributes was evaluated across 16 distinct age groups ranging from early childhood (age 5 years) to advanced age (age 80 years). We employed the nonparametric Wilcoxon signed-rank test and Cliff’s Delta (d) to estimate effect size, with subsequent estimation of 95% confidence intervals. We identified significant deviations from the control group in the ability to perform activities of daily living (ADL), cognitive function, dementia prevalence, and learning capacity.

### Ability to Perform ADL: An Age-Dependent Decline

P value: <0.0019 for all age groups; Average Effect Size: -0.876 +/- 0.064

Interpretation: These data support a large and negative impact across the age spectrum compared to controls. The marked negative effect size seen at ages 5 to 30 years underscores a potential critical age range for early ADL compromise.

### Cognitive Function: Ageing and Potential Early AD Indicators

P value: <0.0019 for all cohorts; Average Effect Size: = -0.895 +/- 0.052

Interpretation: This result indicates a large negative effect size seen at ages 5 to 30 years, potentially signifying early cognitive markers predating the clinical diagnosis of AD.

### Dementia: Stability and Late-life Variation

P value: <0.0019; Average Effect Size: = 0.969 +/- 0.016

Interpretation: Dementia scores are relatively stability with age with a large positive mean effect. The observed large effect begins early (5-30 years) and persists throughout the lifespan of affected subjects.

Learning: Insight into Declining Neuroplasticity with Advancing Age P value: <0.0019; Average Effect Size: = -0.969 +/- 0.016

Interpretation: Learning capabilities were negatively impacted evidenced by an average large effect that highlights the role of learning as a potential marker for early AD detection. Once again, this effect size is apparent at ages 5 to 30 years in this longitudinal study.

### Summary of Findings and Their Implications on AD Research

Collectively, these analyses underscore distinct age-related patterns that correlate with AD progression markers. The large effect sizes seen as early as ages 5 to 30 years and around the age of 50 years emphasizes potential pivotal phases in AD-related decline, warranting further exploration to elucidate mechanistic underpinnings and to fortify early diagnostic frameworks. These results are summarized in Table 1a.

### Psychological Related Feature Effects

#### Anxiety

P value: all <0.0019; Average Effect Size: = 0.822 +/- 0.049

Interpretation: There is a significant deviation in the level of anxiety in the APOE ε3/4 genotype groups compared to the control groups. The positive effect size supports a large difference compared to the control subjects. The largest effect sizes are observed from age 5 to 30 years and again at age 50 years and beyond.

#### Major Depression

P value: <0.0019 except for ages 35-45 years; Average Effect Size: = 0.605 +/- 0.141

Interpretation: The data for major depression was variable across the age cohorts with significant differences observed across most cohorts. The average Cliff’s delta supports an overall medium effect size that peaks beginning at age 50 years and beyond.

The data for the APOE ε3/4 genotype suggests that there are varying trends in psychological factors associated with age. Anxiety effects are increased in early childhood to age 30 years and then increase again at age 50 and beyond, which could represent critical ages for psychological changes. For major depression, there are notable fluctuations at early ages with more stability in later ages, but with an increasing trend as age progresses at 50 years and beyond.

### Neurobiological and Neurodegenerative Biomarkers: (A1-B stands for Amyloid Beta)

For the neurobiological and neurodegenerative biomarkers data, here is the trend analysis from age 5 years onwards:

#### A1-B (non-Amyloidogenic) and A-B1 Plaques

P value: <0.0019; Average Effect Size: = 0.979 +/- 0.013

Interpretation: There is a significant difference in the level of both features in the APOE ε3/4 genotype groups compared to the controls. The large positive effect sizes for both factors begin early and remain consistently large, across all age cohorts.

#### A-B1 (42/40) Protein

P value: <0.0019; Average Effect Size: = 0.974 +/- 0.019

Interpretation: The average effect size for this biomarker was 0.974+/- 0.019 across all but one age cohort but all age cohorts experienced a large positive effect size (i.e., >0.80).

#### A-B1 Oligomers

P value: <0.0019; Average Effect Size: = 0.954 +/- 0.019

Interpretation: There is a significant difference in Amyloid-Beta Oligomers compared to controls. While there is slight variability, the large positive effect size (i.e., >0.9) begins early and persists across all age cohorts.

#### A-B1 (42/40) Clearance

P value: <0.0019 for all cohorts beyond age 40 years; Average Effect Size: = -0.487 +/- 0.284

Interpretation: Throughout the early years, the differences compared to the control groups do not achieve statistical significance except transiently at age 20 years of age. This factor shows some variability in effect size early on and then a sustained decrease at ages beyond 35 years with the average effect size of -0.977+/-0.029 across all subsequent age groups.

#### Tau Protein Aggregation, Tau Protein-P, and Tau Protein/MAPT

P value: <0.0019 for all three features; Average Effect Sizes: >0.950 with 95% CIs between 0.013 and 0.028

Interpretation: All three Tau Protein related factors, Tau Protein Aggregation, Tau Protein-P, and Tau Protein/MAPT were significantly different from their respective control groups. Similarly, effect sizes were consistently large and positive. These changes from controls are observed early and persist across all age matched cohorts.

Overall: The stability of most markers at a value around 0.984 suggests a large positive effect size across all ages, with a notable exception in A-B1 (42/40) Clearance. The decrease in Amyloid-Beta1 (42/40) Clearance from middle-age onwards appears to indicate a pattern associated with aging and the APOE ε3/4 genotype worthy of further research.

### Neuroinflammatory and Oxidative Stress Markers

In the data for neuroinflammatory and oxidative stress markers, we can observe the following trends from age 5 years onwards:

#### Neuroinflammation

P value: <0.0019 for all age groups except ages 35-45 years; Average Effect Sizes: 0.900 +/- 0.049

Interpretation: There are significant differences between the groups from age 5 to 30 years but not from age 35 to 45 years. At 50 years and onwards the significant differences return and persist until the end of the study. Overall, the effect sizes are large and positive even with the medium effect size to a from age 35 to 45 years.

#### Neuronal Oxidative Stress

P value: <0.0019 for all age groups except ages 35-45 years; Average Effect Sizes: 0.856 +/- 0.072

Interpretation: There are significant differences between the groups from age 5 to 30 years, but these differences fail to achieve statistical significance between age 35 to 45 years. At 50 years and onwards the significant differences return and persist until the end of the study. Overall, there was a large positive average effect size. The nadir of effect sizes between age 35 and 45 years was 0.617+/-0.075 (medium) before returning to a relatively large and consistent effect size.

#### PGRN (Progranulin)

P value: <0.0019 observed at ages 20, 30, 60, 65, and 80 years; Average Effect Sizes: -0.261 +/- 0.227

Interpretation: There are sporadic statistically significant differences between the groups. Regarding effect size, this protein shows considerable variability, with a notable decrease in effect size between ages 15 and 20 years (i.e., +0.039 to -0.905). Overall, the average effect size was negative and variably small.

#### PGRN-Brain

P value: <0.0019 observed at ages 20-30 years only; Average Effect Sizes: -0.276 +/- 0.214

Interpretation: Statistically significant differences were only observed at ages 20 to 30 years. During this same age range the effect size becomes large and negative (-0.932+/-0.051). Overall, the effect size was small and negative except for the period 20 to 30 years of age.

These trends indicate a relatively consistent and large effect size for the APOE ε3/4 genotype on Neuroinflammation and Neuronal Oxidative stress except for a brief slight decrease between ages 35 and 45 years. For PGRN and PGRN-Brain the results are more variable with a notable decrease in effects size for PGRN-Brain at ages 20 to 40 years.

These results could be indicative of the body’s response to oxidative stress and inflammation, which are factors related to aging and neurodegenerative processes such as Alzheimer’s disease. The story for PGRN and PGRN-Brain in AD remains incomplete. However, further research and analysis is needed to determine the significance and effect sizes of these trends.

#### Neuronal Integrity and Damage Indicators

The trends for neuronal integrity and damage indicators from age 5 onwards are as follows:

#### Brain Atrophy

P value: <0.0019 for all age groups; Average Effect Sizes: 0.969 +/- 0.214

Interpretation: Statistically significant differences between control and affected groups were observed across all age cohorts. The effect size is also consistently large and positive with all effect values are >0.900.

#### Normal Motor Function

P value: <0.0019 for all age groups except ages 5-10 years; Average Effect Sizes: -0.782 +/- 0.065

Interpretation: Analysis of the youngest age groups did not reveal significant differences between the groups. From age 15 years on the differences in deviations from normal motor function become statistically significant. Overall, the average effect size was large and negative. There is a variable medium to large negative effect sizes for age 5 to 20 years (-0.512 to -0.905). Small negative effect sizes are observed at ages 25 to 30 years. At age 40 years and beyond the effect size becomes large and negative (-0.964+/-0.065).

#### Neurofilament-Light (NFL)

P value: <0.0019 for all age groups; Average Effect Sizes: 0.954 +/- 0.024

Interpretation: This Neuroinflammation marker was significantly higher in the APOE ε3/4 genotype groups for all age groups studied. The importance of this protein is reinforced by the consistently large and positive average effect size.

These trends suggest consistent levels of increased brain atrophy throughout the age range of 5 to 80 years, with only minor fluctuations. N Motor Function shows considerable variability, with significant declines indicating worsening motor function with age. Neurofilament Light (NFL) remains relatively stable, suggesting consistently large effect sizes for this marker across different ages, with perhaps minor decreases in mid-life. These patterns may indicate the natural progression of neuronal integrity and damage as age increases, potentially reflecting the underlying neurobiological changes associated with aging in the context of the APOE ε3/4 genotype.

#### Quality of Life Indicators

The trends for Quality-of-Life indicators related to physical and psychological aspects from age 5 years and onwards are as follows:

#### QofL-Physical

P value: <0.0019 for all age groups except 35-45 years; Average Effect Sizes: - 0.817 +/- 0.063

Interpretation: The differences between the control and affected groups were statistically significant for all age groups except for the 35 to 45 years cohorts. The effect sizes show some variability but on average were large and negative. The 35 to 45 cohorts experienced a medium and negative effect size of -0.643+/-0.051.

#### QofL-Psychological

P value: <0.0019 for all age groups except 35-45 years; Average Effect Sizes: - 0.822 +/- 0.067

Interpretation: These results mirror the data for physical quality of life. The average effect sizes were also large and negative. The 35 to 45 years cohorts experienced some improvement but still experienced an average medium and negative effect size of -0.617+/-0.051 during this period.

These trends suggest a general pattern of some fluctuation in the quality-of-life indicators, with modest improvements and deteriorations at different life stages. The significant decrease at age 50 for both physical and psychological aspects could indicate a critical period of challenge in quality of life, potentially related to life events, health status changes, or other factors.

#### Behavioral and Social Measures

The trends for behavioral and social measures from age 5 years onwards are as follows:

#### Abnormal Behavior

P value: <0.0019 for all age groups; Average Effect Sizes: -0.891 +/- 0.051

Interpretation: Abnormal behavioral issues in the affected cohorts were significantly different from controls in all groups from 5 to 80 years of age. The average effect size across all cohorts was large and positive. This feature remained at a constant near maximum effects size of 0.984 through childhood until age 30, after which a slight improvement was observed between ages 35 and 45 years. At age 50 and beyond, the average effect size was large and positive with slight variability at 65 to 70 years.

#### Sleep Disturbance

P value: <0.0019 for all age groups; Average Effect Sizes: -0.891 +/- 0.051

Interpretation: Difficulties with sleeping in the affected cohorts were also significantly different from controls in all age groups, Similarly to the abnormal behavior issues, sleep disturbance effect size was initially constant at >0.900 and improves slightly from age 35 to 45 years. The effect size increased again at age 50 with an average Cliff’s d of 0.860+/- 0.058, with small fluctuations before reaching a maximum effect size 0.984 at age 80 years.

#### Social Behavior

P value: <0.0019 for all age groups at and beyond age 50 years; Average Effect Sizes: -0.667 +/- 0.196

Interpretation: Social behavior in the affected groups was variably different from the control groups. The differences were statistically significant from ages 5 to 15 years, again at age 25 and then from age 50 years on. The average effect size was negative and medium. It began with a large negative value < -0.800 and did not change until age 20, before improving substantially at age 25, followed by a substantial decrease again at age 30. After some variability in the ensuing years, it decreased and stabilized beyond age 50 at a large negative value (-0.888+/-0.027).

#### Speech & Language

P value: <0.0019 for all age groups beyond age 35 years; Average Effect Sizes: -0.812+/-0.153

Interpretation: p values fail to achieve significance at age 5 and from 25 to 35 years of age. There was a statistically significant difference at all ages beyond age 35 years. The effect size was small for age 5 years. There was a dramatic decrease from -0.360 at age 5 to -0.984 at age 20. Effect size improved slightly at age 25 to 30 (effect size was small at - 0.118 for both groups). Overall, the effect size was negative and large. At age 35 and beyond the effect size remained large and negative with an average value of -0.975+/-0.013 .

#### The Diagnosis of Alzheimer’s disease (AD)

P value: <0.0019 for all age cohorts; Average Effect Sizes: 0.973+/-0.013

Interpretation: The effect of the APOE ε3/4 genotype on the Dx of AD was evaluated in the same fashion as the 27 features above that are associated with the risk of developing AD and disease progression. The difference between the control group and the groups with the APOE ε3/4 genotype were statistically significant in all age cohorts. The average effect was large and positive. Individual effect sizes never fell below 0.900, consistent with a sustained large positive effect of the 27 features on an AD diagnosis.

These observed trends suggest that while abnormal behavior and sleep disturbance experienced some fluctuations, they tended to return to baseline values by age 80. In contrast, social behavior, and speech & language showed deterioration over time, with social behavior displaying notable variability and speech & language decreasing to and remaining at the lowest score from mid-life onwards. These patterns might reflect the progressive impact of aging in the context of APOE ε3/4 mutations on behavioral and social functions. Finally, the 27 input features produced a large and positive effect on the Dx of AD beginning at an early age. Further research will be required to determine the true significance and magnitude of these trends.

### RESULTS for APOE ε4/4 Genotype vs Controls (see graphic summary in Table 2.)

**Table 2:**
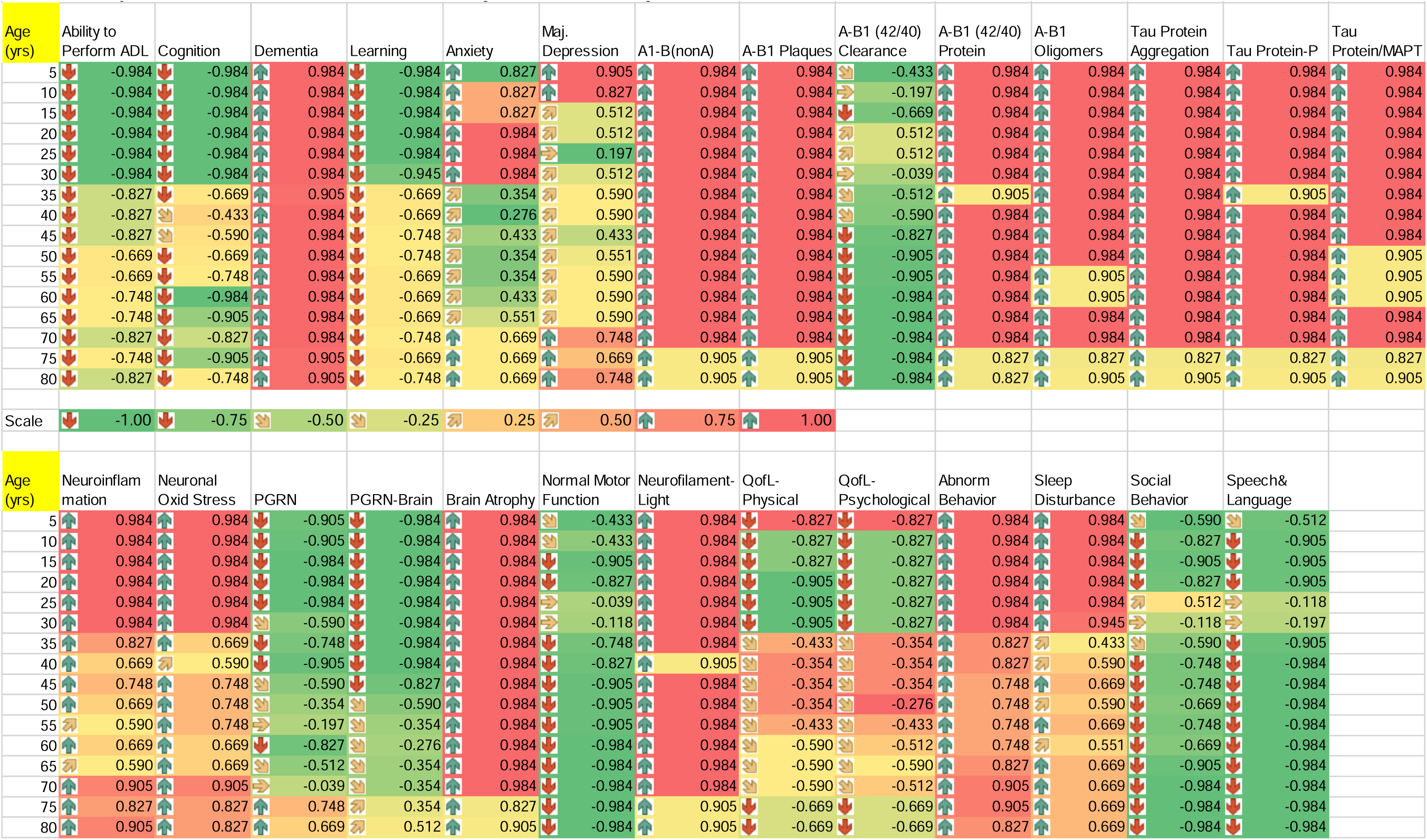
Summary of the Cliff’s delta effect size data for the APOE ε4/4 gene variant across all Age cohorts.

APOE ε4/4 Genotype: Alterations in Cognitive and Functional Metrics Across Age Span Reflect Potential AD Progression Markers

### Statistical Examination of Longitudinal Changes in AD-Related Features

The longitudinal trajectory of Alzheimer’s Disease (AD)-related cognitive and functional features was studied across 16 distinct age groups ranging from early childhood (age 5 years) to advanced age (age 80 years). We employed the nonparametric Wilcoxon signed-rank test and Cliff’s Delta (d) with the Hedges correction of 0.984 for the sample size of 25 pairs per cohort to estimate true effect sizes, with their 95% confidence intervals. We i focused on identifying significant deviations from control groups in the ability to perform activities of daily living (ADL), cognitive function, dementia, and learning capacity.

### Ability to Perform ADL: An Age-Dependent Decline

P value: <0.0019 for all cohorts; Average Effect Size: -0.851 +/- 0.058)

Interpretation: These data support a large and negative impact across the age spectrum from 5 to 45 years of age compared to controls. From age 50 on there was some variability but the effect size was at least medium with an average effect size of -0.748 +/-0.076. The marked negative effect size seen at ages 5 to 45 years underscores a broad potential critical age range for early ADL compromise.

### Cognitive Function: Ageing and Potential Early AD Indicators

P value: <0.0019 for all cohorts; Average Effect Size: = -0.836 +/- 0.086

Interpretation: These results indicate a large negative effect size seen at ages 5 to 30 years, potentially signifying early cognitive markers pertinent to AD onset. In midlife there was a medium negative effect size (-0.682 +/- 0.105). After age 55 years the effects sizes remain negative and large (-0.874 +/- 0.079).

### Dementia: Stability and Late-life Variation

P value: <0.0019 for all cohorts; Average Effect Size: = 0.969 +/- 0.016

Interpretation: Dementia scores are relatively consistent across all age groups with a large positive average effect. The observed large effect begins early and persists throughout the lifespan of affected subjects.

### Learning: Insight into Declining Neuroplasticity with Advancing Age

P value: <0.0019; Average Effect Size: = -0.804 +/- 0.067

Interpretation: Learning capabilities were significantly negatively impacted by the APOE ε4/4 genotype evidenced by an average large effect that highlights the role of learning as a potential marker for early AD detection. Once again, this effect size was apparent early, beginning at ages 5 to age 30 years in this longitudinal study. Beyond age 30 years a medium negative effect size persists (-0.701 +/- 0.025).

### Summary of Findings and Their Implications on AD Research

Collectively, these data underscore distinct age-related patterns that correlate with AD progression markers. The large effect sizes seen as early as ages 5-30 years and around the age of 50 years emphasizes a potential pivotal phase in AD-related decline, warranting further exploration to elucidate mechanistic underpinnings and to support early diagnostic frameworks.

### Psychological Related Feature Effects

#### Anxiety

P value: <0.0019 for age groups 5 to 30 years and beyond 65 years of age only; Average Effect Size: = 0.637 +/- 0.122

Interpretation: There was a significant deviation in the level of anxiety in the APOE ε4/4 genotype groups compared to the control groups at specific ages. The positive effect sizes support a large difference compared to the control subjects. The largest effect sizes are observed from age 5 to 30 years and again at age 65 years and beyond.

#### Major Depression

P value: <0.0019 except for ages 20-30 years; Average Effect Size: = 0.598 +/- 0.081

Interpretation: The data for major depression was variable across the age cohorts with significant differences observed across early and late cohorts. The average effect size supports a medium effect size. The effect size peaks from age 5 to 10 years. and again after 65 years.

These data suggest that there are varying trends in psychological factors associated with age. Anxiety effects increased in early childhood to age 30 years and then increase again at age 65 years and beyond, which could represent critical ages for AD related psychological changes. For major depression, there are notable fluctuations at early ages with more stability in later ages, but with an increasing trend as age progresses at 65 years and beyond.

### Neurobiological and Neurodegenerative Biomarkers: (A1-B stands for Amyloid Beta)

For the neurobiological and neurodegenerative biomarkers data, here was the trend analysis from age 5 years onwards:

#### A1-B (non-Amyloidogenic) and A-B1 Plaques

P value: <0.0019 for both features across all age cohorts; Average Effect Size: = 0.979 +/- 0.013 for both features

Interpretation: There was a significant difference in the level of both features in the APOE ε4/4 genotype groups compared to controls. The large effect sizes for both factors begin early and remain consistently large, across all age cohorts.

#### A-B1 (42/40) Protein

P value: <0.0019 across all cohorts; Average Effect Size: = 0.959 +/- 0.027

Interpretation: The average effect size for this biomarker was large and positive across all age groups.

#### A-B1 Oligomers

P value: <0.0019 across all cohorts; Average Effect Size: = 0.959 +/- 0.023

Interpretation: There was a significant difference in Amyloid-Beta Oligomers in affected subjects compared to controls. While there was minimal variability in the effect size, the large positive effect size (i.e., >0.9) begins early and persists across all age cohorts.

#### A-B1 (42/40) Clearance

P value: <0.0019 for all cohorts beyond age 30 years; Average Effect Size: = -0.561 +/- 0.251

Interpretation: Throughout the early years, the differences compared to the control groups do not achieve statistical significance. This factor shows some variability in effect size during the early years and then a steady decrease at ages beyond 35 years with the average effect size of -0.945+/-0.041 across all subsequent age groups.

#### Tau Protein Aggregation, Tau Protein-P, and Tau Protein/MAPT

P value: <0.0019 for all three features; Average Effect Sizes: >0.950 with 95% CIs between 0.021 and 0.024

Interpretation: All three Tau Protein related factors, Tau Protein Aggregation, Tau Protein-P, and Tau Protein/MAPT were significantly different from their respective control groups. Similarly, effect sizes were consistently large and positive. Importantly, these changes from controls are observed early and persist across all age matched cohorts.

#### Overall

The pattern of most markers suggests a large positive effect size across all ages, with a notable exception in A-B1 (42/40) Clearance. The persistent large decrease in Amyloid-Beta1 (42/40) Clearance from early childhood onwards appears to indicate a clear pattern associated with aging in the context of the APOE ε4/4 genotype that demands further research.

### Neuroinflammatory and Oxidative Stress Markers

In the data for neuroinflammatory and oxidative stress markers, we can observe the following trends from age 5 years onwards:

#### Neuroinflammation

P value: <0.0019 for all age groups; Average Effect Sizes: 0.831 +/- 0.075

Interpretation: There are statistically significant differences between the affected and control groups at all ages studied. Overall, the average effect size was positive and large even considering a noticeable decrease in effect size to 0.656+/-0.047 (medium) occurring from age 40 to 65 years.

#### Neuronal Oxidative Stress

P value: <0.0019 for all age groups; Average Effect Sizes: 0.831 +/- 0.070

Interpretation: There were statistically significant differences between the affected and control groups at all ages studied. Overall, the average effect size was positive and large even including a noticeable decrease in effect size to 0.695+/-0.051 (medium) occurring from age 35 to 65 years.

#### PGRN (Progranulin)

P value: <0.0019 observed at ages 5-45, 60-65, and 75-80 years only; Average Effect Sizes: -0.507 +/- 0.272

Interpretation: There are sporadic statistically significant differences between the groups. Regarding effect size, this protein shows considerable variability, with a large negative effect size between ages 5 and 25 years (-0.953+/-0.038). Overall, the average effect size at age 30 years and beyond was negative and variably small to medium.

#### PGRN-Brain

P value: <0.0019 observed at ages 5 to 50 years; Average Effect Sizes: -0.610 +/- 0.242

Interpretation: Statistically significant differences were observed from age 5 to 50 years. Differences did not achieve significance in any cohort beyond age 50 years. During the earliest age range the effect size was large and negative (- 0.967+/-0.034). Over the age range of 55 to 70 years affected subjects experienced a small negative effect size (-0.386+/- 0.078). In the final decade of 75 to 80 years the effects size became small and positive (0.433+/-0.154). Overall, the effect size was medium and negative.

These trends indicate a relatively consistent and large effect size of the APOE ε4/4 genotype on Neuroinflammation and Neuronal Oxidative stress except for a brief slight decrease between ages 35 and 45 years. For PGRN and PGRN-Brain the results are more variable with a notable large and negative effects size for PGRN-Brain at ages 5to 45 years.

These results could be indicative of the body’s response to inflammation and oxidative stress, which are factors related to aging and neurodegenerative processes such as Alzheimer’s disease. The story for PGRN and PGRN-Brain in AD remains incomplete. However, further research was needed to determine the significance and of the large and negative effect sizes of these trends. observed as early as 5 years of age and continuing through midlife.

#### Neuronal Integrity and Damage Indicators

The trends for neuronal integrity and damage indicators from age 5 onwards are as follows:

#### Brain Atrophy

P value: <0.0019 for all age groups; Average Effect Sizes: 0.969 +/- 0.021

Interpretation: Statistically significant differences between control and affected groups were observed across all age cohorts. The effect size was also consistently large and positive with all values > 0.800.

#### N Motor Function

P value: <0.0019 for age groups 15-20 years and then 40 to 80 years; Average Effect Sizes: -0.748 +/- 0.155

Interpretation: Analysis of the youngest two age groups did not reveal significant differences between the groups. From age 15 to 20 years and again from 40 years onward, the differences in deviations from normal motor function were statistically significant. Overall, the average effect size was borderline large and negative. There was a variable small to large negative effect sizes for age 5 to 30 years (-0.118 to -0.905). At age 40 years and beyond the effect size became large and negative (-0.940+/-0.037).

#### Neurofilament-Light (NFL)

P value: <0.0019 for all age groups; Average Effect Sizes: 0.969 +/- 0.016

Interpretation: This Neuroinflammation marker was significantly higher in the APOE ε4/4 genotype groups for all age group cohorts studied. The importance of this protein was reinforced by the consistently large and positive average effect sizes.

These trends suggest consistent levels of increased brain atrophy throughout the age range of 5 to 80 years, with only minor fluctuations. Normal Motor Function shows considerable variability, with significant declines indicating worsening motor function with age. Neurofilament Light (NFL) remains relatively stable, suggesting consistently large effect sizes for this marker across different ages, with perhaps minor decreases in mid-life. These patterns may indicate the natural progression of neuronal integrity and damage as age increases, potentially reflecting the underlying neurobiological changes associated with aging in the context of the APOE ε4/4 genotype.

#### Quality of Life Indicators

The trends for Quality-of-Life indicators related to physical and psychological aspects from age 5 years and onwards are as follows:

#### QofL-Physical

P value: <0.0019 for age groups 5 to 30 years and again from 70 to 80 years; Average Effect Sizes: - 0.640 +/- 0.102

Interpretation: The differences between the control and affected groups did not achieve statistical significance for cohorts from 35 to 65 years. The effect sizes show some variability but on average were medium and negative. The affected age groups 5 to 30 years experienced a large negative effect size (-0.866 +/- 0.035). The remaining age cohorts experience an average medium effect size of -0.540 +/- 0.081.

#### QofL-Psychological

P value: <0.0019 for all age groups except 35-45 years; Average Effect Sizes: - 0.602 +/- 0.102

Interpretation: These results mirror the data for physical quality of life. The effect sizes overall were also medium and negative. The 5 to 30 years cohorts also experience large and negative effect sizes (-0.827 +/- 0.000). The remaining age cohorts experience on average a medium effect size of -0.505 +/- 0.087.

These trends suggest a general pattern of modest improvements and deteriorations at different life stages with some fluctuations in the quality-of-life indicators. The large negative effect sizes at age 5 to 30 years for both physical and psychological aspects could indicate a critical period of challenge in quality of life that demands further investigations.

#### Behavioral and Social Measures

The trends for behavioral and social measures from age 5 years onwards were as follows:

#### Abnormal Behavior

P value: <0.0019 for all age groups; Average Effect Sizes: 0.876 +/- 0.049

Interpretation: Abnormal behavioral issues in the affected cohorts were significantly different from controls in all groups from 5 to 80 years of age. The average effect size across cohorts was large and positive until age 45 to 60 years when there was some evidence of minimal improvement. This feature remains at a constant near maximum effects size of 0.984 through childhood to age 30 years of age. At age 65 years and beyond, the average effect size remains large and positive.

#### Sleep Disturbance

P value: <0.0019 for all age groups; Average Effect Sizes: 0.753 +/- 0.093

Interpretation: Similarly to the abnormal behavior issues, sleep disturbance effect sizes were large and positive from ages 5 to 30 years at 0.977+/- 0.013. The effect size becomes more variable at age 35 and beyond with an average of 0.618 +/- 0.049, consistent with a medium positive effect.

#### Social Behavior

P value: <0.0019 for age groups at ages between 10 and 25 years. A significant difference between the APOE ε4/4 and control groups from age 40 to 80 years.; Average Effect Sizes: -0.674 +/- 0.187

Interpretation: Overall, the average effect size was negative and medium. It began with a medium negative value at age 5 years but between ages 10 and 20 years became large. From age 25 to 65 years the effect size was variable and medium (-0.472+/- 0.311.From age 65 years on the average effect size was large and negative (-0.964 +/- 0.039).

#### Speech & Language

P value: <0.0019 for age groups 10 to 20 years and from age 35 to 80 years; Average Effect Sizes: -0.831+/-0.141

Interpretation: p values failed to achieve significance at age 5 and then from 25 to 30 years of age. There was a statistically significant difference at all ages at and beyond age 35 years. The effect size was medium for age 5 years (- 0.512). The effect size was transiently small (-0.157 +/- 0.077) for the cohorts aged 25- and 30-year-old. At age 35 and beyond the effect size remained large and negative with an average effect size of -0.976+/-0.015 .

#### The Diagnosis of Alzheimer’s disease (AD)

P value: <0.0019 for all age cohorts; Average Effect Sizes: 0.984+/-0.000

Interpretation: The effect of the APOE ε4/4 genotype on the Dx of AD was evaluated in the same fashion as the 27 features above that were associated with the risk of developing AD and disease progression. The average effect size on the Dx of AD was large. positive and sustained.

These observed trends suggest that while abnormal behavior and sleep disturbance experience some fluctuations, they tend to return to large effect levels over time. In contrast, social behavior, and speech & language show deterioration over time, with social behavior displaying notable variability and speech & language decreasing to and remaining at the lowest score from mid-life onwards. These patterns could reflect the progressive impact of aging on behavioral and social functions in the context of the APOE ε4/4 genotype. Finally, the 27 input features produce a large and positive effect on the 28^th^ feature namely the Dx of AD beginning at an early age.

## Discussion

The intricate relationship between APOE genotypes and Alzheimer’s Disease (AD) progression has been a cornerstone of genetic research within the AD community. Through the application of the aiHumanoid platform, our study delves into the varied progression patterns associated with APOE ε3/4 and APOE ε4/4 genotypes, uncovering novel biomarkers and genotypic changes indicative of early-stage AD.

### APOE Genotypes and AD Progression

The seminal work by Corder et al. (1993) established the APOE ε4 allele as a significant risk factor for AD, a finding that has been pivotal in guiding subsequent research [2]. Genin et al. (2011) further elucidated the semi-dominant inheritance of the APOE ε4 allele, emphasizing its critical role in AD’s genetic landscape [3]. Our findings, revealing early biomarkers and phenotypic changes in individuals aged 5 to 30 years, represent a significant departure from traditional AD research which has focused predominantly on older adults. This shift underscores the potential for early diagnostic interventions that could alter the disease’s trajectory, highlighting the critical need for new screening protocols tailored to younger populations at risk. Recent advancements in biomarker research, such as the multicohort study of serum phospho-tau181, emphasize the potential for these markers in early diagnosis of AD (Ashton et al., 2021) [31].

Recent neuropathological evidence from Serrano-Pozo et al. (2023) corroborates the significant impact of the APOE ε4 allele on amyloid accumulation in the brain, reinforcing the allele’s role in AD’s progression [24]. This aligns with our findings, which highlight early biomarkers specific to APOE ε3/4 and APOE ε4/4 genotypes, potentially paving the way for earlier diagnosis and intervention strategies. Jansen et al. (2023) also contributes to this story by demonstrating the negative cognitive implications of cerebral amyloid-β aggregation, further underscoring the clinical significance of genotype-specific research [25].

### Biomarker Advancements and Early Detection

The pursuit of reliable biomarkers for early AD detection has been significantly advanced by the work of Zetterberg and Blennow (2021), who discuss the status and prospects of AD biomarkers [26]. Their insights into the biomarker landscape provide a crucial context for our study’s identification of novel biomarkers associated with APOE genotypes, suggesting a pathway toward non-invasive, early diagnostic protocols.

### Cellular Mechanisms and Therapeutic Targets

De Strooper and Karran (2022) offered a comprehensive overview of the cellular processes in AD, delving into the molecular mechanisms underlying disease progression [27]. This framework is vital for the interpretation of our findings regarding phenotypic alterations, suggesting that these cellular changes could facilitate the development of targeted therapies. Furthermore, Cummings et al. (2021) emphasize the critical role of precision medicine in the AD drug development pipeline, emphasizing the importance of biomarker-guided therapeutic strategies [28].

The genotype-specific changes observed through aiHumanoid simulations offer promising pathways for the development of targeted therapeutic interventions. These findings pave the way for precision medicine approaches in AD, where treatments can be tailored based on an individual’s genetic and phenotypic risk profile, potentially long before the onset of clinical symptoms. A recent review on the AD drug development pipeline, including precision medicine approaches and diversity of targeted therapies, reflects the growing emphasis on such strategies (Cummings et al., 2022) [32].

### The Future of AD Research and Clinical Practice

The role of autophagy in AD, as discussed by Takahashi et al. (2022), presents potential therapeutic targets that are congruent with our study’s emphasis on genotype-specific pathways [29]. Additionally, Liu et al. (2013) provide a detailed examination of APOE ’s risk mechanisms and therapeutic potential, underscoring the importance of translating genetic and biomarker research into viable treatment strategies [4].

Our study underscores the need for a bridge between molecular insights and clinical application. The role of AI and technology in advancing precision neurology for AD, including early detection and intervention strategies, is increasingly recognized (Hampel et al., 2021) [33]. This approach aligns with our findings, emphasizing the potential of AI to revolutionize AD research and care.

Furthermore, the ethical and psychological considerations of identifying at-risk individuals at early stages cannot be overlooked. The potential impact of disclosing biomarker information, particularly in younger populations, requires careful navigation to ensure that the benefits of early diagnosis are balanced against the possible risks to the individual (Bemelmans et al., 2016) [34].

The critical importance of longitudinal studies in understanding AD progression and the impact of early interventions has been well-documented (Schneider et al., 2014) [14]. Our research contributes to this body of knowledge, suggesting that monitoring from a young age could be key in developing effective prevention strategies in high-risk populations.

Importantly, it remains to be determined if younger individuals with a convincing family history of dementia and who miss several critical developmental milestones may constitute such a high-risk population.

In conclusion, by unveiling the early indicators of AD risk within younger populations, our research not only enriches the scientific dialogue around AD but also offers a beacon of hope for early intervention strategies. It is incumbent upon us, the scientific and medical communities, to translate these insights into tangible benefits, crafting a future where AD’s burden is significantly reduced through early biomarker detection and personalized care (Jack et al., 2018) [30].

### Limitations of the present study

Our research demonstrates the significant potential of artificial intelligence (AI) in modeling the early diagnosis and progression of Alzheimer’s Disease (AD) using the aiHumanoid platform. Despite its innovative approach, the study faces several limitations:

1. <u>Translatability to Real-World Scenarios:</u> The virtual subject simulations, grounded in extensive medical literature, genetic data, and physician review, may not capture the complete complexity of human biology. The simplification within these simulations might affect the precision of our findings’ applicability to real patient scenarios, particularly concerning the intricate interplay of genetic, environmental, and lifestyle factors in AD.
2. <u>Focus on Genetic Variants:</u> The initial phase of our study focuses solely on two APOE genotypes, overlooking other genetic elements contributing to AD’s heterogeneity. In subsequent parts of this longitudinal study, we plan to explore a broader spectrum of genetic factors and their interactions to achieve a comprehensive understanding of AD.
3. <u>Biological Variability and Disease Evolution:</u> Designed for homogeneity aside from specific mutations, our virtual cohorts may not fully represent the biological variability and spontaneous disease evolution characteristic of more diverse human populations. We intend to mitigate this limitation by expanding the virtual study’s scope to other mutations (APP, PSEN1 and PSEN2) and utilizing real-world data in future research.
4. <u>Inherent Design Limitations:</u> Governed by the assumptions and constraints within AI simulations, our study’s design could limit the identification of more subtle disease markers and the exploration of biomarkers’ therapeutic potential.
5. <u>Ethical Considerations:</u> Although we exclusively used data from hypothetical AI generated subject in the current study, the ethical implications of AI in medical research—such as data privacy, consent, and the interpretation of virtual patient data—were not addressed. Ensuring responsible use of AI technologies in healthcare, especially concerning anonymized real patient data, will be imperative.

Recognizing these potential limitations, this AI directed virtual longitudinal study still offers significant insights into AD’s progression and highlights the transformative potential of AI in both research and clinical management. Future efforts will be dedicated to enhancing AI simulation realism, diversifying the genetic and environmental factors examined, and addressing the ethical challenges posed by the integration of AI in healthcare. Part 2 of this virtual longitudinal study will focus on the Amyloid Precursor Protein (APP), PSEN1 and PSEN2 mutations while an anticipated Part 3 will address combinations of AD associated risk factors.

## Data Availability

All data produced in the present work are contained in the manuscript

## References

1. Caselli, R.J., Dueck, A.C., Osborne, D., et al. (2009). “Longitudinal modeling of age-related memory decline and the APOE ε4 effect.” New England Journal of Medicine, 361(3): 255–263.

2. Corder, E.H., Saunders, A.M., Strittmatter, W.J., et al. (1993). “Gene Dose of Apolipoprotein E Type 4 Allele and the Risk of Alzheimer’s Disease in Late Onset Families.” Science, 261(5123): 921–923.

3. Genin, E., Hannequin, D., Wallon, D., et al. (2011). “APOE and Alzheimer disease: a major gene with semi-dominant inheritance.” Molecular Psychiatry, 16(9): 903–907.

4. Liu, C.C., Kanekiyo, T., Xu, H., Bu, G. (2013). “Apolipoprotein E and Alzheimer disease: risk, mechanisms, and therapy.” Nature Reviews Neurology, 9(2): 106–118.

5. Mahley, R.W., Weisgraber, K.H., Huang, Y. (2006). “Apolipoprotein E: structure determines function, from atherosclerosis to Alzheimer’s disease to AIDS.” Journal of Lipid Research, 50(Supplement): S183–S188.

6. Raulin, AC., Doss, S.V., Trottier, Z.A. et al. ApoE in Alzheimer’s disease: pathophysiology and therapeutic strategies. Mol Neurodegeneration 17, 72 (2022). 10.1186/s13024-022-00574-4

7. Reiman, E.M., Arboleda-Velasquez, J.F., Quiroz, Y.T., et al. (2020). “Exceptionally low likelihood of Alzheimer’s dementia in APOE2 homozygotes from a 5000-person neuropathological study.” Nature Communications, 11(1): 667.

8. Sun, YY., Wang, Z. & Huang, HC. Roles of ApoE4 on the Pathogenesis in Alzheimer’s Disease and the Potential Therapeutic Approaches. Cell Mol Neurobiol 43, 3115–3136 (2023). 10.1007/s10571-023-01365-1

9. APOE C130R (ApoE4)” (December 6, 2022). APOE [R176C];[C130R] (ApoE2/4) | ALZFORUM

10. Bu, G. *APOE* targeting strategy in Alzheimer’s disease: lessons learned from protective variants. Mol Neurodegeneration 17, 51 (2022). 10.1186/s13024-022-00556-6

11. Leggett H, A rare mutation protects against Alzheimer’s disease, Stanford-led research finds” (May 31, 2022). A rare mutation protects against Alzheimer’s disease, Stanford-led research finds | News Center | Stanford Medicine

12. Cummings J, Zhou Y, Lee G, Zhong K, Fonseca J, Cheng F. Alzheimer’s disease drug development pipeline: 2023. Alzheimers Dement (N Y). 2023 May 25;9(2):e12385. doi: 10.1002/trc2.12385. Erratum Alzheimers Dement (N Y). 2023 Jun 28;9(2):e12407.

13. Hampel, H., O’Bryant, S.E., Molinuevo, J.L., Zetterberg, H., Masters, C.L., Lista, S., Kiddle, S.J., Batrla, R., Blennow, K. (2020). Blood-based biomarkers for Alzheimer disease: mapping the road to the clinic. Nature Reviews Neurology, 16(11), 589–600.

14. Schneider, L.S., Mangialasche, F., Andreasen, N., Feldman, H., Giacobini, E., Jones, R., Mantua, V., Mecocci, P., Pani, L., Winblad, B., Kivipelto, M. (2014). Clinical trials and late-stage drug development for Alzheimer’s disease: An appraisal from 1984 to 2014. Journal of Internal Medicine, 275(3), 251–283.

15. Esmail S and Danter WR (2021) NEUBOrg: Artificially Induced Pluripotent Stem Cell-Derived Brain Organoid to Model and Study Genetics of Alzheimer’s Disease Progression. Front. Aging Neurosci. 13:643889. doi: 10.3389/fnagi.2021.643889

16. WR Danter, An aiHumanoid Simulation of Gram-Negative Sepsis: A Comprehensive Multi-Organoid Platform for Advanced Disease Modeling, Drug Discovery, and Personalized Medicine, bioRxiv 2023.07.18.549304; doi: 10.1101/2023.07.18.549304

17. WR Danter Advancing Drug Development with aiHumanoid Simulations: A Virtual Phase 1 Comparative Study of Standard Chemotherapy versus Standard Chemotherapy plus COTI-2 for Pancreatic Adenocarcinoma, medRxiv 2023.09.08.23295256; doi: 10.1101/2023.09.08.23295256

18. WR Danter, HAI-VECT(SCD): AI-Humanoid Enabled Virtual Clinical Trial for Sickle Cell Disease. medRxiv 2023.10.17.23297152; doi: 10.1101/2023.10.17.23297152

19. Sally Esmail, Wayne R Danter, DeepNEU©: Introducing aiCRISPRL, a hybrid AI stem cell and organoid simulation platform with broad gene editing capabilities and applications bioRxiv 2022.06.18.496679; doi: 10.1101/2022.06.18.496679

20. Cliff, N. (1996). Ordinal Methods for Behavioral Data Analysis (1st ed.). Psychology Press. 10.4324/9781315806730

21. Romano, J., Kromrey, J. D., Coraggio, J., & Skowronek, J. (2006) Appropriate statistics for ordinal level data: Should we really be using t-tests and Cohen’s d for evaluating group differences on the NSSE and other surveys? Presented at the Annual Meeting of the Florida Association of Institutional Research

22. Zhiyuan Wan, Xin Xia, David Lo and Gail C. Murphy (2019) How does Machine Learning Change Software Development Practices? IEEE Transactions on Software Engineering. https://www.researchgate.net/publication/335425733

23. Hedges, L. V. (1981). Distribution theory for Glass’s estimator of effect size and related estimators. Journal of Educational Statistics, 6(2), 107–128. doi:10.3102/10769986006002107

24. Serrano-Pozo, A., Qian, J., Monsell, S.E., et al. (2023). “APOEε4 allele and amyloid pathology in the elderly: A postmortem neuropathological study.” Neurology, 100(1), e1–e12.

25. Jansen, W.J., Ossenkoppele, R., Tijms, B.M., et al. (2023). “Association of Cerebral Amyloid-β Aggregation With Cognitive Functioning in Persons Without Dementia.” JAMA Psychiatry, 80(1), 37–47.

26. Zetterberg, H., & Blennow, K. (2021). “Biomarkers for Alzheimer’s Disease: Current Status and Prospects for the Future.” Journal of Internal Medicine, 290(4), 817–836.

27. De Strooper, B., & Karran, E. (2022). “The Cellular Phase of Alzheimer’s Disease.” Cell, 184(1), 32–44.

28. Cummings, J., Lee, G., Ritter, A., Sabbagh, M., & Zhong, K. (2021). “Alzheimer’s disease drug development pipeline: 2021.” Alzheimers & Dementia (N Y), 7(1), e12179.

29. Takahashi, R.H., Capetillo-Zarate, E., & Lin, M.T. (2022). “New insights into the role of autophagy in Alzheimer’s disease.” Journal of Clinical Investigation, 132(2), e143739.

30. Jack, C.R., Bennett, D.A., Blennow, K., et al. (2018). “NIA-AA Research Framework: Toward a biological definition of Alzheimer’s disease.” Alzheimer’s & Dementia.

31. Ashton, N.J., Janelidze, S., Al Khleifat, A., et al. (2021). “A multicohort study of serum phospho-tau181 in Alzheimer’s disease.” Nature Medicine, 27, 1184–1192.

32. Cummings, J., Lee, G., Zhong, K., Fonseca, J., & Taghva, K. (2022). “Alzheimer’s disease drug development pipeline: 2022.” Alzheimer’s & Dementia: Translational Research & Clinical Interventions, 8(1), e12295.

33. Hampel, H., Vergallo, A., Afshar, M., et al. (2021). “Precision neurology for Alzheimer’s disease: A multidisciplinary integrative approach paving the way to precision medicine.” Precision Clinical Medicine, 4(1), 1–15.

34. Bemelmans, S.A.S., Tromp, K., Bunnik, E.M., et al. (2016). “Psychological, behavioral, and social effects of disclosing Alzheimer’s disease biomarkers to research participants: a systematic review.” Alzheimer’s Research & Therapy, 8, 46.

## Reference list for Dx feature list for AD

A1. Ability to Perform ADL (Activities of Daily Living): Sikkes, S.A.M., de Lange-de Klerk, E.S.M., Pijnenburg, Y.A.L., Scheltens, P., & Uitdehaag, B.M.J. (2009). A systematic review of Instrumental Activities of Daily Living scales in dementia: Room for improvement. Journal of Neurology, Neurosurgery, & Psychiatry, 80(1), 7.

A2. Cognition: Jack, C.R., Bennett, D.A., Blennow, K., et al. (2018). NIA-AA Research Framework: Toward a biological definition of Alzheimer’s disease. Alzheimer’s & Dementia, 14(4), 535–562.

A3. Dementia: McKhann, G.M., Knopman, D.S., Chertkow, H., et al. (2011). The diagnosis of dementia due to Alzheimer’s disease: Recommendations from the National Institute on Aging-Alzheimer’s Association workgroups on diagnostic guidelines for Alzheimer’s disease. Alzheimer’s & Dementia, 7(3), 263–269.

A4. Learning: Selkoe, D.J. (2002). “Alzheimer’s Disease is a synaptic failure.” Science.

A5. Anxiety: Gulpers B, Ramakers I, Hamel R, Köhler S, Oude Voshaar R, Verhey F. Anxiety as a Predictor for Cognitive Decline and Dementia: A Systematic Review and Meta-Analysis. Am J Geriatr Psychiatry. 2016 Oct;24(10):823–42. doi: 10.1016/j.jagp.2016.05.015.

A6. Major Depression: Diniz BS, Butters MA, Albert SM, Dew MA, Reynolds CF 3rd. Late-life depression and risk of vascular dementia and Alzheimer’s disease: systematic review and meta-analysis of community-based cohort studies. Br J Psychiatry. 2013 May;202(5):329–35. doi: 10.1192/bjp.bp.112.118307.

A7. A1-B (non-atherogenic): Mahley RW, Weisgraber KH, Huang Y. Apolipoprotein E4: a causative factor and therapeutic target in neuropathology, including Alzheimer’s disease. Proc Natl Acad Sci U S A. 2006 Apr 11;103(15):5644-51. doi: 10.1073/pnas.0600549103.

A8. A-B1 Plaques: Hardy J, Selkoe DJ. The amyloid hypothesis of Alzheimer’s disease: progress and problems on the road to therapeutics. Science. 2002 Jul 19;297(5580):353–6. doi: 10.1126/science.1072994. Erratum in: Science 2002 Sep 27;297(5590):2209

A9. A-B1 (42/40) Clearance: Mawuenyega, K.G., Sigurdson, W., Ovod, V., et al. (2010). “Decreased clearance of CNS β-amyloid in Alzheimer’s disease.” Science vol. 330, no. 6012

A10. A-B1 (42/40) Protein: Bateman, R.J., Xiong, C., Benzinger, T.L.S., et al. (2012). “Clinical and biomarker changes in dominantly inherited Alzheimer’s disease.” N Engl J Med 2012; vol. 367 no. 9:795–804 DOI: 10.1056/NEJMoa1202753

A11. A-B1 Oligomers: Klein, W.L., Krafft, G.A., & Finch, C.E. (2001). “Targeting small Aβ oligomers: The solution to an Alzheimer’s disease conundrum?” Trends in Neurosciences. Volume 24, Issue 4, 1 April 2001, Pages 219–224

A12. Tau Protein Aggregation: Götz, J., Chen, F., van Dorpe, J., & Nitsch, R.M. (2001). “Formation of neurofibrillary tangles in P301l tau transgenic mice induced by Aβ42 fibrils.” Science. 24 Aug 2001 Vol 293, Issue 5534 pp. 1491–1495 <u>DOI:</u> 10.1126/science.1062097

A13. Tau Protein-P (Phosphorylated Tau): Iqbal K, Liu F, Gong CX. Tau and neurodegenerative disease: the story so far. Nat Rev Neurol. 2016 Jan;12(1):15–27. doi: 10.1038/nrneurol.2015.225.

A14. Tau Protein/MAPT (Microtubule-Associated Protein Tau): Lee VM, Goedert M, Trojanowski JQ. Neurodegenerative tauopathies. Annu Rev Neurosci. 2001;24:1121–59. doi: 10.1146/annurev.neuro.24.1.1121.

A15. Neuroinflammation: Heneka MT, Carson MJ, El Khoury J, et al. Neuroinflammation in Alzheimer’s disease. Lancet Neurol. 2015 Apr;14(4):388–405. doi: 10.1016/S1474-4422(15)70016-5.

A16. Neuronal Oxidative Stress: Butterfield DA, Halliwell B. Oxidative stress, dysfunctional glucose metabolism and Alzheimer disease. Nat Rev Neurosci. 2019 Mar;20(3):148–160. doi: 10.1038/s41583-019-0132-6.

A17. PGRN (Progranulin): Finch N, Baker M, Crook R, et al. Plasma progranulin levels predict progranulin mutation status in frontotemporal dementia patients and asymptomatic family members. Brain. 2009 Mar;132(Pt 3):583–91. doi: 10.1093/brain/awn352.

A18. PGRN-Brain: Karamysheva ZN, Tikhonova EB, Karamyshev AL. Granulin in Frontotemporal Lobar Degeneration: Molecular Mechanisms of the Disease. Front Neurosci. 2019 Apr 25;13:395. doi: 10.3389/fnins.2019.00395.

A19. Brain Atrophy: Fox NC, Schott JM. Imaging cerebral atrophy: normal ageing to Alzheimer’s disease. Lancet. 2004 Jan 31;363(9406):392–4. doi: 10.1016/S0140-6736(04)15441-X

A20. Normal Motor Function: Aggarwal NT, Wilson RS, Beck TL, Bienias JL, Bennett DA. Motor dysfunction in mild cognitive impairment and the risk of incident Alzheimer disease. Arch Neurol. 2006 Dec;63(12):1763–9. doi: 10.1001/archneur.63.12.1763.

A21. Neurofilament-Light: Zetterberg, H., Wilson, D., Andreasson, U., et al. Plasma tau levels in Alzheimer’s disease. Alz Res Therapy 5, 9 (2013). 10.1186/alzrt163

A22. QofL-Physical, QofL-Psychological: Logsdon RG, Gibbons LE, McCurry SM, Teri L. Assessing quality of life in older adults with cognitive impairment. Psychosom Med. 2002 May-Jun;64(3):510–9. doi: 10.1097/00006842-200205000-00016.

A23. Abnormal Behavior: Steinberg M, Shao H, Zandi P, et al; Cache County Investigators. Point and 5-year period prevalence of neuropsychiatric symptoms in dementia: the Cache County Study. Int J Geriatr Psychiatry. 2008 Feb;23(2):170–7. doi: 10.1002/gps.1858.

A24. Sleep Disturbance: Ju YE, Lucey BP, Holtzman DM. Sleep and Alzheimer disease pathology--a bidirectional relationship. Nat Rev Neurol. 2014 Feb;10(2):115–9. doi: 10.1038/nrneurol.2013.269.

A25. Social Behavior: Cummings, J. L., Mega, M., Gray, K., Rosenberg-Thompson, S., Carusi, D. A., & Gornbein, J. (1994). The Neuropsychiatric Inventory: Comprehensive assessment of psychopathology in dementia. Neurology, 44(12), 2308–2314.

A26. Speech and Language: Mesulam, M.-M. (2003). “Primary progressive aphasia—A language-based dementia.” New England Journal of Medicine. October 16, vol. 349 no. 16 DOI: 10.1056/NEJMra022435

A27. Diagnosis of AD: (a) Albert, M.S., DeKosky, S.T., Dickson, D., et al. (2011). “The diagnosis of mild cognitive impairment due to Alzheimer’s disease: Recommendations from the National Institute on Aging-Alzheimer’s Association workgroups on diagnostic guidelines for Alzheimer’s disease. “Alzheimer’s & Dementia. 2011;7(3):270–279), (b) Porsteinsson, A.P., Isaacson, R.S., Knox, S. et al. Diagnosis of Early Alzheimer’s Disease: Clinical Practice in 2021. J Prev Alzheimers Dis 8, 371–386 (2021). 10.14283/jpad.2021.23

## Reviews

A28. Jack, C.R. Jr., Bennett, D.A., Blennow, K., et al. (2018). “NIA-AA Research Framework: Toward a biological definition of Alzheimer’s disease.” Alzheimer’s & Dementia.

A29. Hampel H, O’Bryant SE, Durrleman S, et al; Alzheimer Precision Medicine Initiative. A Precision Medicine Initiative for Alzheimer’s disease: the road ahead to biomarker-guided integrative disease modeling. Climacteric. 2017 Apr;20(2):107–118. doi: 10.1080/13697137.2017.1287866.

A30. Ballard C, Gauthier S, Corbett A, Brayne C, Aarsland D, Jones E. Alzheimer’s disease. Lancet. 2011 Mar 19;377(9770):1019–31. doi: 10.1016/S0140-6736(10)61349-9.

A31. Selkoe DJ, Hardy J. The amyloid hypothesis of Alzheimer’s disease at 25 years. EMBO Mol Med. 2016 Jun 1;8(6):595–608. doi: 10.15252/emmm.201606210.

A32. Heneka MT, Carson MJ, El Khoury J, et al. Neuroinflammation in Alzheimer’s disease. Lancet Neurol. 2015 Apr;14(4):388–405. doi: 10.1016/S1474-4422(15)70016-5.

